# Circumcision for HIV prevention in men who have sex with men: an updated meta-analysis

**DOI:** 10.1101/2025.07.02.25330772

**Authors:** Stephanie M. Davis, Ian Fellows, Oscar de Leon, Parsa Bastani, Avi J Hakim, Carlos Toledo, Ugonna Ijeoma, John Schneider, Todd Lucas, Megan Peck, David Philpott

## Abstract

**Background:** While circumcision reduces HIV risk for men who have sex with women, its value for men who have sex with men (MSM) remains unclear. Any protection would be for primarily-insertive MSM (PI-MSM) and could have substantial impact given high incidence. Prior meta-analyses were ambiguous due to inconsistent, highly-confounded data, leaving the total weight of evidence unclear even after a small 2024 trial in PI-MSM showed protection.

**Setting:** No geographic or other setting restrictions were used.

**Methods:** We performed an updated literature search for data on the association of male circumcision with HIV among MSM. We conducted descriptive analyses and a random-effects meta-analysis to assess the main association among PI-MSM. To isolate the protective effect of circumcision from potential confounding factors, we also performed a confounder-adjusted meta-analysis, comparing the effect of circumcision on HIV status among primarily-insertive MSM versus among other (non-primarily-insertive) MSM in studies with stratified results.

**Results:** Forty-nine studies were included in the descriptive analysis, 13 in the unadjusted meta-analysis, and 10 in the confounder-adjusted meta-analysis. In the unadjusted meta-analysis, circumcision was associated with a lower risk for HIV among PI-MSM (odds ratio (OR) = 0.57, 95% CI: 0.33-0.98); publication bias was present. In the confounder-adjusted analysis, the additional effect of circumcision in the PI-MSM group – a ratio of ORs - was 0.53 (95% CI: 0.34– 0.83), indicating a lower bound on the protective effect of circumcision among PI-MSM adjusting for confounding factors. Publication bias was not present.

**Conclusions:** Among PI-MSM, circumcision was protective against HIV. Findings support male circumcision as an effective HIV prevention method for PI-MSM, alongside established combination prevention methods including pre-exposure prophylaxis.

## Background

Circumcision reduces men’s risk of acquiring HIV through sex with women by approximately 60%.^1,2,3^ Since 2007, the World Health Organization (WHO) has recommended voluntary medical male circumcision (VMMC) for HIV prevention in countries with generalized HIV epidemics and low circumcision coverage^4^. Through 2023, fifteen sub-Saharan African countries had provided over 37 million circumcisions to men seeking protection from HIV^5^, averting nearly 900,000 infections during that period and 1.4 million by 2030.^6^

However, for gay, bisexual, and other men who have sex with men (MSM) – who account for 34% of all new HIV infections outside SSA annually^7^ – the picture has been less clear. Data are plentiful but inconsistent. Two 2019 meta-analyses^8,9^ (the last of several, including an inconclusive 2011 Cochrane review) noted that data from SSA and Asia generally support protection whereas data from the United States and Europe generally do not. The underlying reasons for these differences are not yet fully understood; confounding is a likely driver in settings where circumcision at birth is associated, positively or negatively, with socioeconomic status and other risk determinants. Another barrier to interpreting the observational data is that because circumcision would directly protect only the insertive partner in any encounter, any effect would be visible only in primarily-insertive (PI)-MSM, and inclusion of MSM preferring receptive or versatile roles would likely dilute the measured effect. In attempting to address this, one of the 2019 meta-analyses found significant protection for PI-MSM and the other did not^8,9^. Finally, data on MSM has other reasons for inconsistency: as a population without a defined sampling frame - unlike general populations in household samples - MSM have often been studied through convenience sampling in unrepresentative venues like sexually transmitted infection clinics.

Faced with these challenges, current WHO recommendations for VMMC (updated 2020) do not include MSM, and recommendations for HIV prevention in MSM (updated 2022)^10^ do not include VMMC. In practice, though MSM seeking VMMC are not excluded or required to disclose their sexual practices, with rare exceptions^11^ they are also not prioritized for demand creation, which has been essential for uptake of this preventive intervention among men who have sex with women.

In 2024, the first randomized trial of VMMC for MSM^12^ reported significant protection at one year among 247 PI-MSM in China. Because there were only five seroconversions, though - all in the control group - and a larger-than-expected effect size (over 90% risk reduction), it is unclear if this small trial is enough to resolve the inconsistency in the prior observational data. Yet, additional trials may be unlikely: they could require infeasibly large sizes if offering pre-exposure prophylaxis (PrEP) medications, which is becoming ethically necessary as their availability broadens.

Clarifying the existing evidence could therefore have important effects. If circumcision protects PI-MSM from HIV, even without expanding VMMC programs to new regions, VMMC could be offered as a prevention option for PI-MSM seeking to reduce their risk for HIV. This would be especially relevant for those facing barriers to long-term use of PrEP and other prevention interventions, who could benefit from ongoing protection without the need for drug adherence or dependence on a reliable health supply chain. Conversely, if VMMC is not protective, limited program resources should not be used to identify men who will not benefit from this intervention, and MSM can be counseled on other evidenced-based approaches that are more protective against HIV.

We consolidated the current evidence by performing 1) an updated systematic review to capture any data added since the 2019 meta-analyses, 2) a standard meta-analysis of effect among PI-MSM, and 3) a novel meta-analysis designed to mitigate confounding and publication bias by using a ratio of odds ratios as its main metric: the comparison between circumcision effects in PI-MSM vs. in other MSM within the same set of studies.

## Methods

### Search strategy

We identified studies in five ways:

1. We assessed all articles included in the 2019 meta-analyses^8,9^ or a 2022 systematic review^13^.
2. We conducted an updated literature search (details in online appendix 1) in Academic Search Complete, Pubmed, Embase, PsychInfo, Cochrane, OVID Global Health, WHO Index Medicus and Scopus for variations of the terms “circumcision”, “MSM” and “HIV” during January 1, 2018--October 16, 2024.
3. We manually searched International AIDS Society (IAS), Conference on Retroviruses and Opportunistic Infections, and HIV Research for Prevention (HIVR4P) abstract archives from 2018 through 2024 for abstracts with the same terms above.
4. We searched biobehavioral surveys (BBSs) among MSM in a CDC-maintained repository. These surveys typically use respondent-driven sampling, assess HIV prevalence, collect risk and behavioral data, and estimate population size. CDC’s repository includes a subset which are CDC-supported. We attempted to acquire additional BBS reports but were unsuccessful.
5. We considered our own primary work shared at IAS 2025.^14^

### Analysis strategy

We planned a descriptive analysis of all observational studies reporting on the association between circumcision and HIV among MSM, followed by the two meta-analyses described above. This strategy was chosen because the two methods had complementary strengths: the former included more datasets as it did not require findings among other (non-PI) MSM, where the latter mitigated bias and confounding by comparing findings by sex role within the same study.

### Inclusion and exclusion criteria

One author (SD) performed screening. Products (articles or abstracts) reporting primary data on any HIV outcome (prevalent or incident infection, including self-reported positive result history) with any observational study design using any metric (odds ratio, hazard ratio, etc.) were includable. Exclusion criteria are shown in table 1.

**Table 1:** Exclusion criteria for descriptive analysis of the association of circumcision with HIV among all MSM and two meta-analyses of this association among.

| Exclusion criterion | Rationale | Descriptive analysis | Unadjusted meta-analysis | Confounder-adjusted meta-analysis |
| --- | --- | --- | --- | --- |
| Duplication, including reporting on the same dataset as another included product.* | Avoids over-weighting datasets with more publications | x | x | x |
| Reporting data from a VMMC-implementing country after substantial scaleup of the VMMC program (>20,000 VMMCs). | VMMC is popularly known in these settings as an HIV prevention tool, so although men with HIV are not excluded from it, if they are less likely to seek it, measured associations become subject to self-selection bias, inflating the apparent protective effect. | x | x | (Counters dataset-wide bias by comparing results between PI-MSM and other MSM within the same study.) |
| Reporting only non-observational data (the RCT) | Allows descriptive analysis to represent the observational data for easy comparison with the trial results, while the other analyses synthesize all available evidence. | x |  | (However, RCT excluded anyway by final criterion below) |
| Not reporting results for a PI-MSM subgroup | Method requires these data |  | x | x |
| Not reporting results for other MSM (non-PI) MSM | Method requires these data |  |  | x |

\* In these situations, we prioritized manuscripts over abstracts, more inclusive and more highly-stratified results, and those representing longer follow-up.

Products were not excluded for being available only through a prior meta-analysis as abstracted data (e.g., dissertations not retrievable by us).

### Data abstraction

One author (SD) abstracted measures of association for all studies in the descriptive analysis. We standardized coefficients of association (CoAs) so that values <1 represent lower risk of HIV infection with circumcision and values >1 represent higher risk. We used adjusted CoAs measures where available; if no CoA was available, we calculated an odds ratio from cell counts. CoAs were also categorized as “lower-risk, significant” (LRS; CoA < 0.94, p<.05), “lower-risk, not significant” (LRNS; p≥.05), “neutral” (CoA 0.94-1.06), “higher-risk, not significant” (HRNS; CoA>1.1), or “higher risk, significant” (HRS).

Other fields abstracted included number of participants, country and region, years of data collection, study design, sampling and recruitment approach, variables adjusted for in calculating the CoA, and stratifications provided (by sex role, and also by whether participants had sex with men only or with men and women). Study design was based on circumcision as the exposure of interest (e.g., an RCT of another intervention was termed cross-sectional if only baseline data were used, or prospective cohort if prospective data was used.)

After studies were identified for inclusion in the meta-analyses, initial abstraction of stratum-specific CoAs and cell counts for PI-MSM and other MSM was repeated by another author (DP) and discrepancies reconciled through discussion. For studies which collected data by sex role but did not report stratum-specific CoAs, we contacted authors to request the additional data needed for the confounder-adjusted meta-analysis (CoAs or cell counts in other MSM).

### Risk of bias assessment

We did not assess risk of bias on studies already included in the prior meta-analyses, but reviewed their risk-of-bias assessments. We assessed newly-identified included studies using the Newcastle-Ottawa scale for prospective cohort studies^15^ and a modified version for cross-sectional studies.^16^

### Data analysis

We performed descriptive analysis on the full set of studies in Excel, including tabulating the distribution of results both by number of studies and by number of participants.

We then completed the unadjusted random-effects meta-analysis in R using the metafor package, estimating pooled log odds ratios (logORs) and their sampling variances among PI-MSM only. We converted effect measures to logORs by log-transforming published odds ratios with their variances derived from reported 95% confidence intervals; we likewise converted risk ratios, hazard ratios, and prevalence ratios to logORs using the generic inverse-variance method. Where an adjusted estimate was unavailable, 2 × 2 contingency tables were used to compute logORs, replacing zero cells with 0.01 to allow estimation. We selected adjusted effect estimates when available and used cell-count estimates otherwise. Finally, we assessed the robustness of findings via two sensitivity analyses: i) a cell-count-only analysis, which excluded studies without raw counts, and ii) a prefer-cell-count analysis, which used cell-count estimates when available and adjusted estimates otherwise.

All models were fitted with restricted maximum likelihood (REML) and a random intercept for study, because several studies contributed multiple strata. The forest plot was ordered by region and estimate variance.

We then conducted what we termed a “confounder-adjusted” meta-analysis, based on the following rationale: Circumcision status is influenced, in childhood and adulthood, by a range of social and personal factors that may also be associated with HIV risk, confounding the analysis and potentially driving the data inconsistencies already discussed. These factors may include:

1. Social or religious group: Certain groups commonly engage in infant, child or adolescent circumcision (e.g., Jewish religion, some ethnic groups). Membership in these groups could be associated - variably across populations - with adult risk behaviors, and/or with assortive sexual mixing causing differential partner pool HIV prevalence.
2. VMMC as risk mitigation: As noted in Table 1, individuals who circumcise to reduce HIV risk are by definition HIV-negative at that time, leading to lower initial HIV prevalence in the group, introducing self-selection bias. This bias would, to our knowledge, be confined to sub-Saharan African countries implementing VMMC, in the absence of large-scale circumcision uptake for HIV prevention elsewhere.

To address these confounders and incorporate as much data as possible, we conducted the confounder-adjusted meta-analysis using the same rules described above but estimating the pooled difference of logORs between PI-MSM and other MSM, among studies providing the needed stratification. The reasoning was that while in a purely non-insertive individual there may be no effect of circumcision on HIV risk, they may have lower or higher HIV prevalence than non-circumcised MSM due to the confounding effects above. If those affect HIV risk in insertive and non-insertive individuals in the same way, then we could “subtract” them out by subtracting the log odds ratio among purely non-insertive MSM from the log odds ratio among insertive individuals. Stated another way, we computed the ratio of the odds ratios.

We considered this a conservative approach with respect to the effect of circumcision on HIV risk from insertive sex, because in reality most MSM do not neatly fall into exclusively insertive or non-insertive categories. Consequently, the studies that stratified by role often classify individuals engaging in some insertive sex into the “least-insertive” category of the stratification. The effect is to dilute the true effect of circumcision on risk and thus we would a priori expect the true protective effect for insertive sex to be stronger than the estimate from the ratio of ratios. A mathematical framework describing how this adjusts for confounding effects is provided in online appendix 2.

For both meta-analyses, publication bias was assessed with funnel plots with asymmetry testing (for all studies, and by dichotomized region group) and estimate heterogeneity testing.

Finally, we repeated the confounder-adjusted analysis dropping the de Leon^14^ et al dataset, which was our own work and the only one drawn from a VMMC country after substantial VMMC scaleup. Code to reproduce the analysis is available in online appendix 2.

## Results

### Included data

For the descriptive analysis, forty-nine observational products representing unique datasets were includable^17,18,19,20,21,22,23,24,25,26,27,28,29,30,31,32,33,34,35,36,37,38,39,40,41,42,43,,44,45, 46,47,48,49, 50,51,52,53,54,55,56,57,58,59,60,61,62,63, 64^ with nine additional products excluded due to criterion 2 only (inclusion flowchart in figure 1).^65,66,67,68,69,70,71,72,73^ Of includable products, five were not included in the prior meta-analyses (two BBSs^22,40^ and three peer-reviewed manuscripts^28,58,62^); all products excluded by criterion 2 only were also new. Missingness was minimal on most variables, primarily occurring in studies available only as abstracted data,^25,34,26,35^, none of which met inclusion criteria for the meta-analyses. Included studies and those excluded based on location (Table 1, criterion 2) are provided in appendix 3. Additional results are in appendix 4.

**Figure 1:**
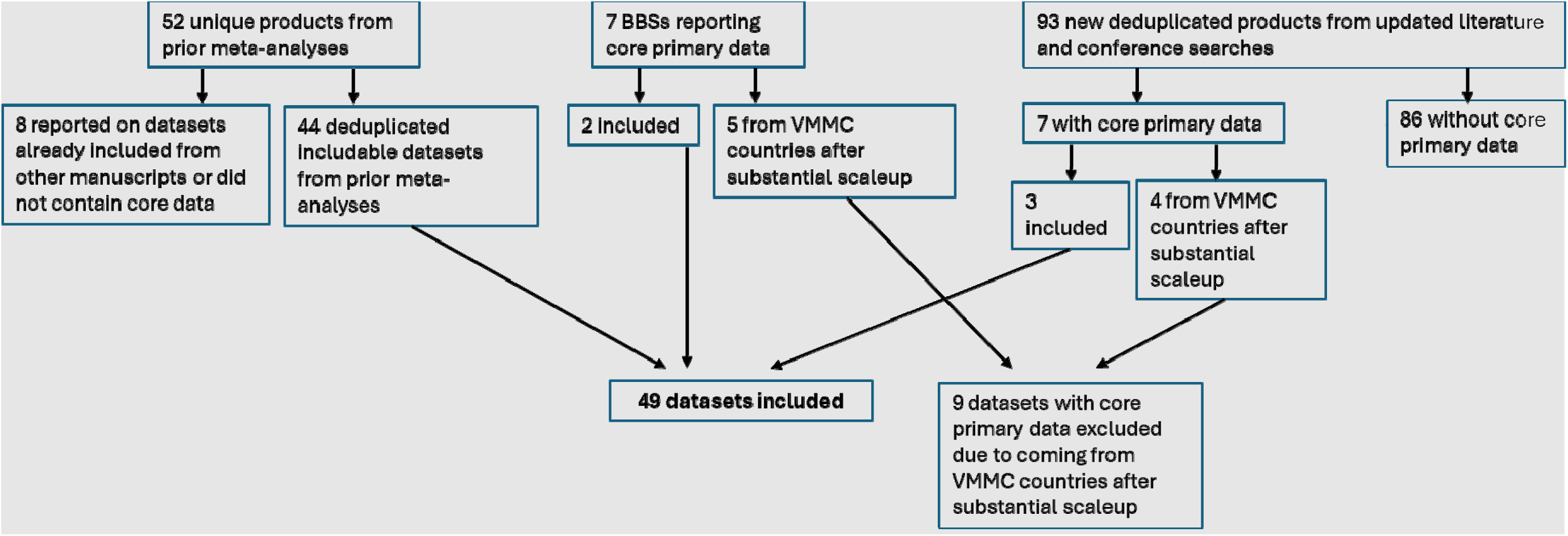
Dataset exclusion flowchart for association of circumcision with HIV among men who have sex with men.

For the unadjusted meta-analysis, 12/49 products were includable, totaling 13 with the trial. (Vs. eight and eleven in the Yuan and Zheng meta-analyses respectively). Definitions for PI-MSM were heterogeneous, encompassing self-report of both preferences and actual practices; time windows from 2-12 months; and cutoffs from ‘no receptive role’ to ‘<50% receptive’, with variable inclusion of “unprotected” as a modifier.

For the confounder-adjusted meta-analysis, eight of the 12 observational products included in the unadjusted meta-analysis were includable, after obtaining additional data from three products’ authors. Also includable were the 2024 trial and our own work (de Leon et al^14^), for a total of 10.

### Descriptive analysis

The 49 observational products were from five major geographic regions (North America, South America and the Caribbean, Europe, Asia/Pacific, sub-Saharan Africa) which we re-sorted for some analyses into US/Canada/Europe/Australia and Asia/Africa/Latin America.

Eight studies were LRS, 23 LRNS, 10 neutral, 7 HRNS, and 1 HRS (figure 2a). This distribution supports a protective association, but when measured by total participant N instead, this pattern is greatly attenuated (figure 2b). The median CoA was odds ratio (OR)=0.79; among studies with significant results, it was OR=0.46, indicating lower odds of HIV with circumcision. Protective associations were commonly seen in studies conducted in Latin America and Asia (14/17; remainder neutral) compared to the United States, Europe and Australia (11/23 protective); and in smaller studies (13/16 among studies with N≤500) compared to larger ones (10/26 among those with N>500). Among the 12/49 studies reporting on PI-MSM – all retrievable manuscripts or abstracts - nine assessed significance: 1 LRS, 4 LRNS, 2 neutral or mixed LRNS/HRNS (stratified by race), 2 HRNS. The median CoA was OR=0.61, again indicating lower odds of HIV with circumcision.

**Figure 2:**
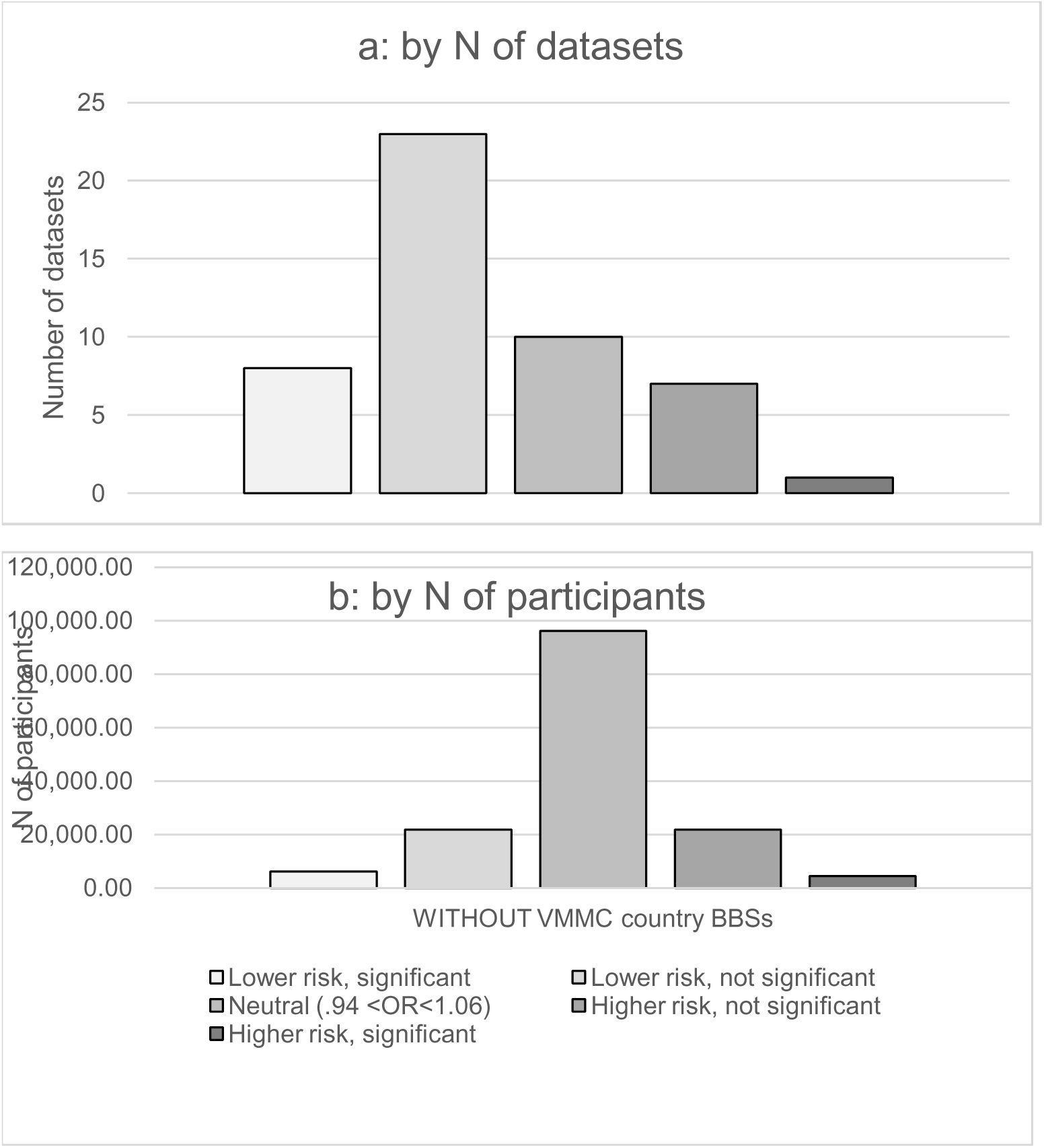
Histogram: datasets on association of circumcision with HIV among men who have sex with men, by category of direction and significance.

### Risk of intra-study bias

Our review of the prior meta-analyses’ assessments of risk for bias in included studies found that Yuan et al rated 52% of their included studies (comprising 33/49 of our included studies) as low-risk for bias, and the remainder high-risk. Zhang et al did not report assessments. Our own bias assessment in the five new included studies found that 4 (cross-sectional) scored below 7/10 (high-risk); one (prospective) scored 8/9 (low-risk). Details are in Appendix 5.

### Unadjusted meta-analysis

This returned an odds ratio (OR) of 0.57 (95% CI 0.33-0.98) for HIV infection in circumcised vs. uncircumcised MSM. (figure 3). The test for heterogeneity returned p=0.02, supporting effect heterogeneity, and the test for asymmetry returned p<0.0001, supporting publication bias. Secondary analyses using cell counts only and preferring cell counts where available returned ORs of 0.56 (0.32-0.98) and 0.62 (0.37-1.03) respectively. Effect differences by region are visible in the forest plot; tests for funnel plot asymmetry returned p<0.01 for both region groups, demonstrating publication bias in both. The effect of region was not significant (p=0.15), but the difference in point estimates was large (logOR-ratio -1.07 for Asia/Africa/Latin America vs. -0.21 for US/Canada/Europe/Australia).

**Figure 3:**
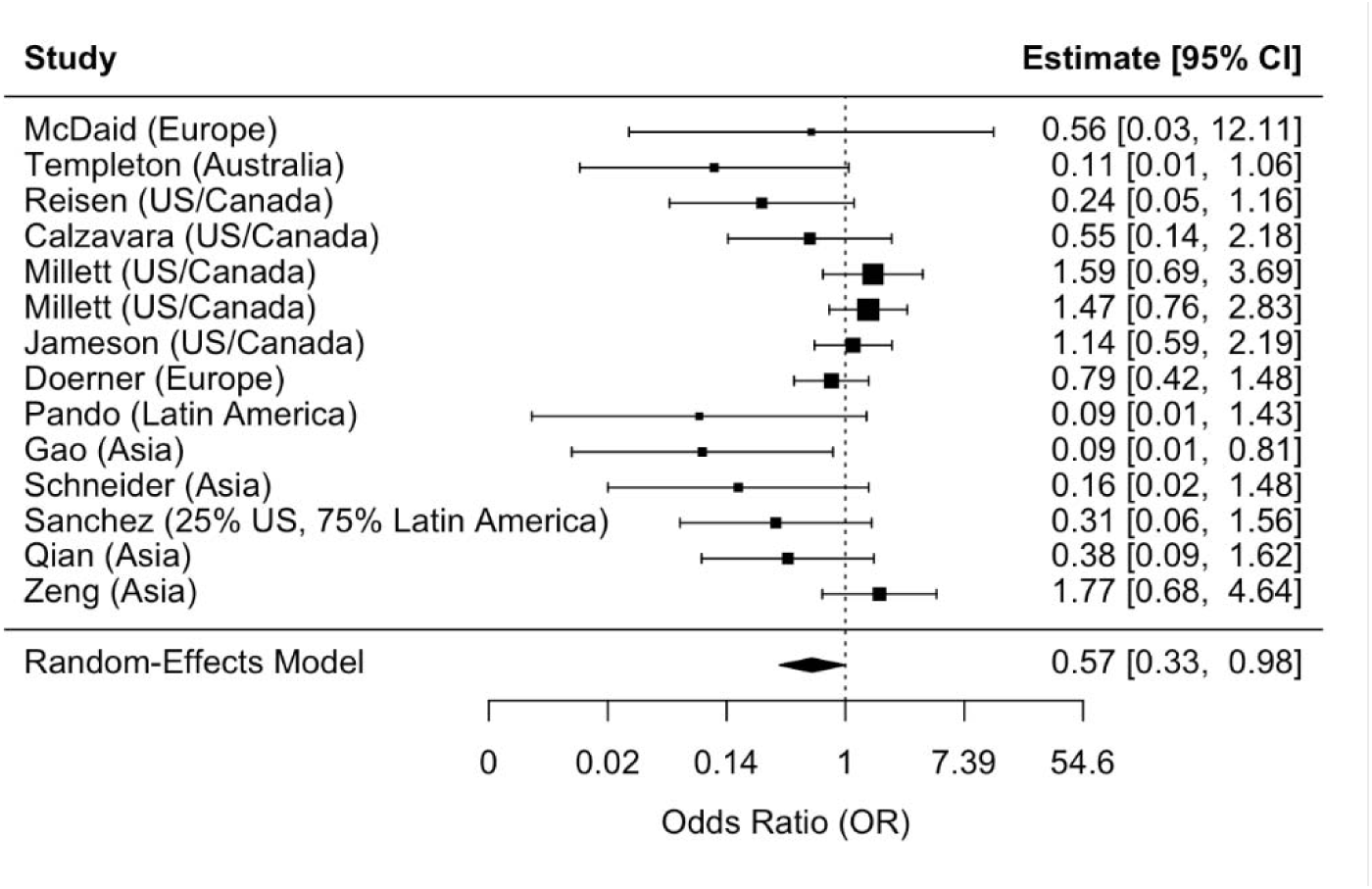
Forest plot, unadjusted meta-analysis of association of circumcision with HIV among primarily-insertive men who have sex with men.

### Confounder-adjusted meta-analysis

This returned a ratio of ratios of 0.53 (p = 0.006; 95% CI 0.34-0.83), indicating a significant and large protective association (forest plot, figure. 4). The test for funnel plot asymmetry returned p = 0.34 (funnel plot, figure 5) and the test for heterogeneity returned p = 0.31, indicating neither publication bias nor heterogeneity on this outcome. Differences by region group were again not significant (p=0.31), this time with a small point estimate for effect of region (logOR-ratio -0.77 vs -0.54). Dropping de Leon et al did not impact the estimate (ratio of ratios = -0.54) or test for heterogeneity; it did impact significance (p = 0.055).

**Figure 4:**
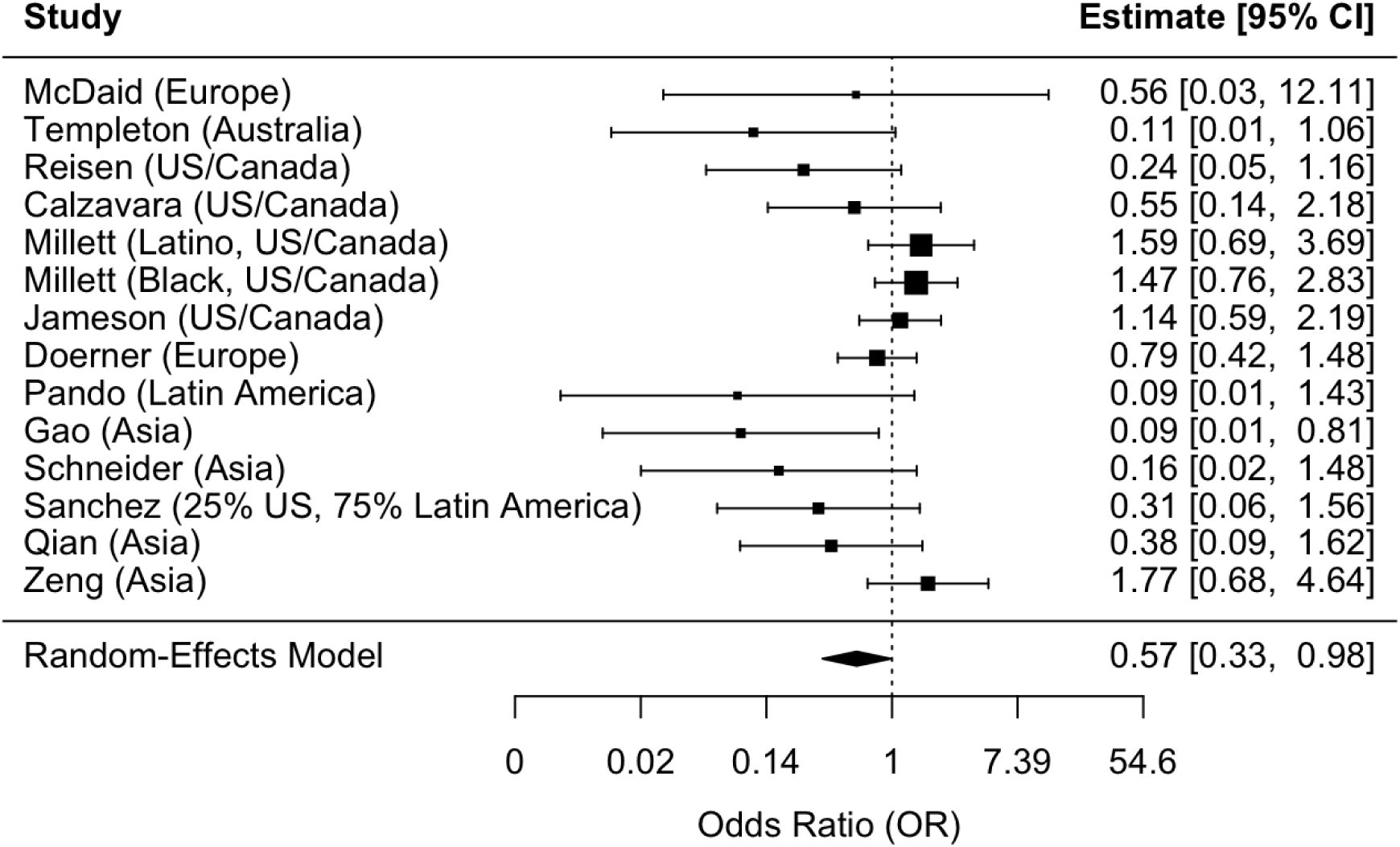
Forest plot, confounder-adjusted meta-analysis (ratio of associations of circumcision with HIV, comparing primarily-insertive MSM to other MSM)

**Figure 5:**
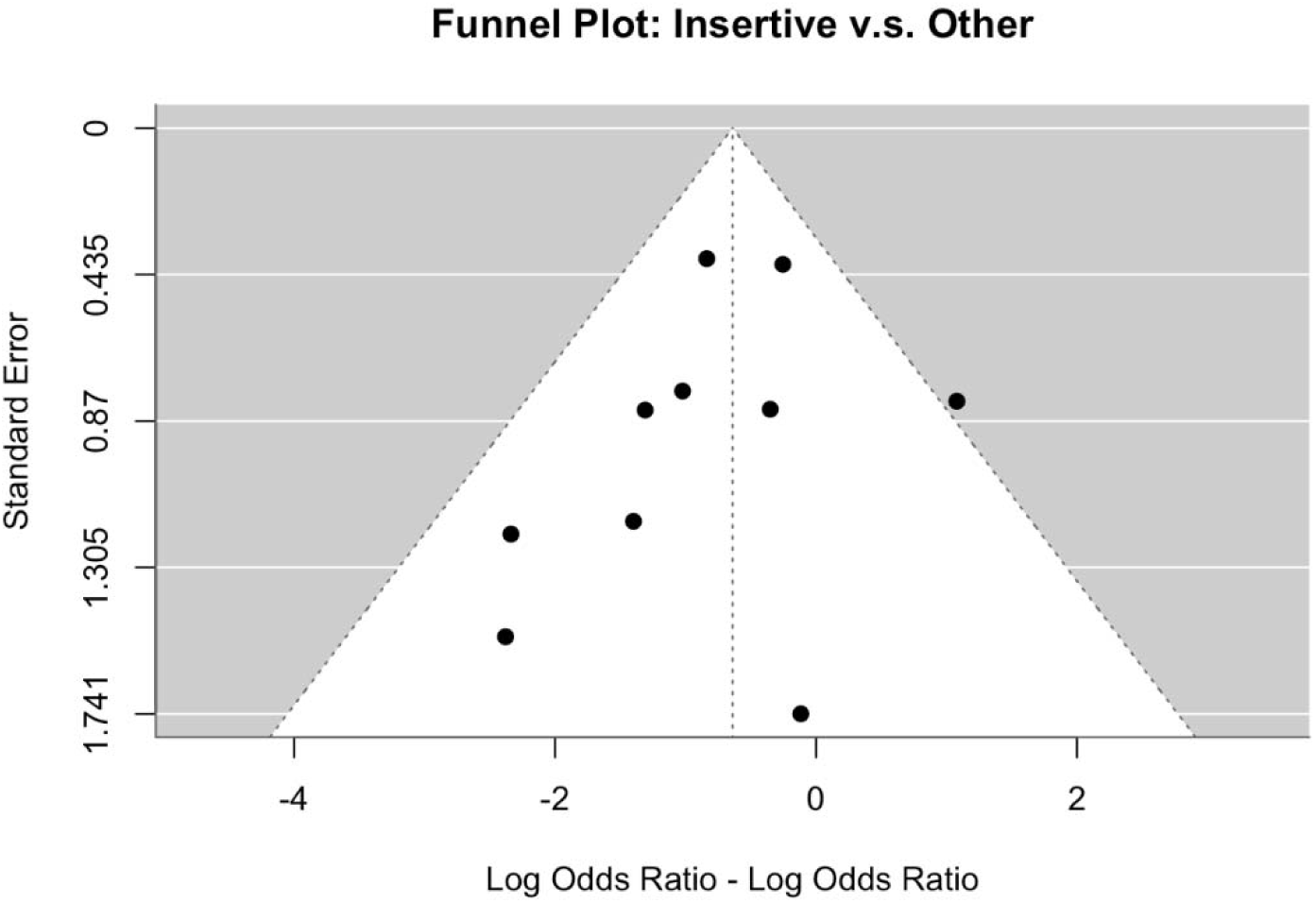
Funnel plot, confounder-adjusted meta-analysis (difference in log odds ratios for associations of circumcision with HIV among men who have sex with men, comparing primarily-insertive MSM to other MSM)

## Discussion

The results of these meta-analyses, along with the 2024 trial, provide new evidence that circumcision protects PI-MSM from HIV acquisition. These findings broaden the potential relevance of VMMC from a single region to global MSM populations. Many settings already have local adolescent/adult medical circumcision services available but have had no reason to seek them.

We do not minimize the remaining scientific challenges. We would expect continuing questions about the basis for offering VMMC to PI-MSM for HIV prevention in the specific regions where data have not suggested effect. Data that could clarify this issue already exist: the United States, Europe and Australia have multiple large MSM cohort registries which could be used in analyses similar to our confounder-adjusted approach to support or challenge our results. They also have past and ongoing HIV prevention trials ripe for similar secondary analyses, in which MSM without HIV were randomized on other interventions but would constitute a prospective cohort on circumcision status.^i^ Small sample sizes for the PI-MSM subgroup in which protective effect is plausible may still require pooling across datasets. In other areas (Latin America, non-VMMC-implementing sub-Saharan African countries), existing cross-sectional BBS datasets often capture circumcision status and sexual positioning; secondary analyses of these could broaden the geographic range of evidence. Either or both approaches would also address what we view as a remaining weakness in the body of evidence: the possibility of residual publication bias not fully mitigated by the analytic approaches taken here. They would also be more feasible and rapid than a second randomized trial.

But we believe based on the balance of the current evidence that it is far more plausible that circumcision has a true protective effect in PI-MSM than that it does not, for these reasons:

1. *Trial evidence*: While small, the single trial was in our view well-conducted, and its findings were clear.
2. *Biological plausibility*: In our view, it is more plausible that the protective effect already established for a penis in the transmission milieu of a vagina would apply in the milieu of an anus than that it would not. While insertive anal sex has a higher per-act transmission risk than insertive vaginal sex^74^, transmission is still too rare (138 transmission/100,000 acts) to overwhelm a substantial protective effect. It is also more plausible that the biological protective effect of circumcision in MSM would be geographically universal, under the existing understanding of its mechanisms^75^ in men who have sex with women, than that regional differences represent true differences.^ii^
3. *Overall observational evidence as shown in our findings*: We found significant protection with circumcision in PI-MSM in both our cofounder-adjusted and unadjusted analyses. Given the lack of purely insertive and non-insertive subgroups in the studies, our adjusted estimates should in fact underestimate this protection for insertive sex, for the reasons described in Methods.^iii^ The common reliance on self-report for circumcision status and sexual role would also be expected to create crossover bias toward the null in all analyses. The inclusion of CDC’s available set of BBSs should also help mitigate bias in the overall body of evidence: these would be free of publication bias on the association of interest, as BBS data are not collected with circumcision as a key outcome and are reliably reported regardless of findings. Intra-study biases are a separate concern, and we and Yuan et al both found the majority of studies were highly susceptible, even if the directionality is unpredictable. However, we propose that our confounder-adjusted analysis mitigates these biases as well, in any situation where they can be assumed to affect PI-MSM and other MSM equally.
4. *Explanations for regional variation*: Known confounders like risk behaviors, and “diluters” like lack of stratification by sex role, are adequate to hide true effects in observational studies; most of the 49 included studies had one or more of these limitations. This situation echoes the evolution of the evidence base for VMMC for men who have sex with women, for whom the initial Cochrane review^76^ discovered ambiguous forest plots rife with publication bias, and declined to rule on efficacy until three randomized trials placed it beyond doubt. Our analyses further support the theory that regional differences reflect uncorrected and/or unmeasured confounding: they are visible in the descriptive analysis, not significant in the unadjusted meta-analysis, and not significant and much smaller in the confounder-adjusted analysis.

The other major scientific question is what proportion of sexual acts must be insertive to qualify as “primarily insertive”. An ideal definition would reflect a true behavioral cutpoint for benefit and would be easy for individual MSM to compare their own practices against. If this is achievable, it would greatly facilitate effective implementation. The heterogeneity of cutpoints used in PI-MSM studies in our work supports the robustness of effect on one hand, but makes it challenging on the other to decide what definition to use for further analyses, let alone what message to give to potential clients. A further consideration is that MSM who seroposition, or use on-demand PrEP or condoms only for receptive sex, would presumably benefit from circumcision similarly to “true” PI-MSM, and might be conceptualized as “functionally PI” or “PI from a risk perspective”, but these nuances are not always captured. In our view, rarity of unprotected receptive anal sex is the most important factor; since its per-unprotected-act infection risk is over 12 times higher^74^ than insertive sex, its attributable risk would be expected to dominate total risk for a given individual once it exceeded 8% of all his unprotected sex acts. That ratio might be one metric worth testing, potentially using sexual diaries for accuracy.

These analyses have strengths and limitations. This is the most current and comprehensive synthesis of the evidence, and the first to our knowledge to incorporate the trial findings. The confounder-adjusted meta-analysis approach seems to have successfully mitigated the sources of bias that have plagued the question to date, and we believe that in the unadjusted meta-analysis, excluding most data from VMMC-implementing countries was another novel conservative measure. For these reasons, and the others discussed above, the true magnitude of protection is likely to exceed our estimates.

However, our analysis, while it supports confounding as the driver of inconsistency between studies – and particularly between regions - does not clarify what the most important confounders may be. Possibilities, which could be tested in some existing primary datasets, include differences in access to PrEP or testing (facilitating risk-mitigating, effect-diluting practices like seropositioning), and in sex role versatility.

We also chose not to include available raw datasets, as they could not meet key quality standards: analysis by authors familiar with the datasets, and some degree of peer review. Conversely, we chose for completeness to include in the descriptive analysis the few datasets we could only access in the prior meta-analyses, though none of these qualified for our meta-analyses. Our search also did not capture regional conferences and was an update search from 2018 onwards, relying on the prior meta-analyses for the earlier literature.

## Conclusions

In our view, while additional analyses of existing datasets could add nuance and strength, the existing evidence indicates that circumcision partially protects PI-MSM from acquiring HIV. This finding could have particular relevance for the millions of such MSM who lack access to injectable PrEP and cannot or do not wish to take oral PrEP effectively.

## Data Availability

All data included in the present study are available in the supplemental materials.

## Data-sharing statement

Data used in these analyses are previously reported, other than the data obtained directly from authors, and are available in the online appendices.

## Acknowledgements

We gratefully acknowledge the contributions of Liviana Calzavara, Sandra Bullock, Julie Riddell, John Schneider, and Lisa McDaid.

## Disclaimer

The views expressed in this manuscript are those of the authors and do not necessarily reflect the official policies of the US Centers for Disease Control and Prevention.

### Online appendices

1. Literature search strategies
2. Meta-analysis code and input values
3. Included articles and articles excluded due to coming from VMMC regions after substantial scaleup – abstracted data
4. Supplementary results: included data and descriptive analysis
5. Risk of bias assessments for new included articles

## Footnotes

i Based on N, capturing circumcision status, and numbers of seroconversions, excellent candidates could include: MOSAICO (HVTN 706), iPrEX, STEP and its extension (HVTN 502), and HPTN 075. (Some include data from VMMC-implementing countries but would be protected from the self-selection bias problem by virtue of being prospective designs.) Other candidates, especially for pooled analysis, could include the Amsterdam Cohort Studies, PROUD, ANRS Prevenir, and IPERGAY.

ii This assertion could be tested by studies comparing the penile microbiomes of circumcised men whose insertive sex practices are with men vs. women, to determine whether the circumcision-induced microbiome shift away from anaerobic, transmission-facilitating bacteria in men who have sex with women applies equally in MSM. Similarly, the presumed universality of biological effect could be challenged by data showing variation across geographic regions in prevalence of anaerobic inflammatogenic bacteria under foreskins in uncircumcised men. We are not aware of such data.

iii Another possible source of bias toward the null is retaining studies that adjusted for STI outcomes. Where these included penile STIs, if circumcision is protective for these in MSM as it is in men who have sex with women, these adjustments introduced collider bias toward the null for HIV outcomes.

