## Appendices for "Circumcision for HIV prevention in men who have sex with men: an updated meta-analysis": appendix 1 circumcision lit review methods.docx

**Requested by: Stephanie Davis**

**Search Query:** circumcision as an HIV prevention method for men who have sex with men (MSM).

**Search Strategy:**

* Searched for variations of terms for “circumcision”, “MSM” and “HIV”; included anything that reported HIV outcomes in both circumcised and uncircumcised MSM; databases = Academic Search Complete, Pubmed, Embase, PsychInfo, Cochrane, OVID Global Health, Scopus)

Probably abstracts went as far back as 2018ish?

| **Database** | **Strategy** | **Run Date** | **Records** |
| --- | --- | --- | --- |
| **Academic Search Complete** | S10 S3 AND S6 AND S9  S9 S7 OR S8  S8 DE "CIRCUMCISION" OR TI (circumcis*) OR AB (circumcis*) OR SU (circumcis*)  S7 DE "CIRCUMCISION"  S6 S4 OR S5  S5 ( (DE "MEN who have sex with men") OR (DE "GAY people") ) OR TI ( (MSM or "men who have sex with men" or gay* or bisexual* or homosexual* or non-heterosexual*). ) OR AB ( (MSM or "men who have sex with men" or gay* or bisexual* or homosexual* or non-heterosexual*) ) OR SU ( (MSM or "men who have sex with men" or gay* or bisexual* or homosexual* or non-heterosexual*) )  S4 (DE "MEN who have sex with men") OR (DE "GAY people")  S3 S1 OR S2  S2 TI ( (Human immunodeficiency virus* or HIV* or AIDS or Acquired Immunodeficiency syndrome*) ) OR AB ( (Human immunodeficiency virus* or HIV* or AIDS or Acquired Immunodeficiency syndrome*) ) OR SU ( (Human immunodeficiency virus* or HIV* or AIDS or Acquired Immunodeficiency syndrome*) )  S1 ((DE "HIV") OR (DE "HIV infections")) OR (DE "AIDS") | 10/16/24 | 18  -18  Duplicates  =0  Unique items |
| **Embase**  **(OVID)**  **1974-** | Embase <1988 to 2024 Week 41>  1 Human immunodeficiency virus/ 137784  2 Human immunodeficiency virus infection/ 319702  3 acquired immune deficiency syndrome/ 133038  4 (Human immunodeficiency virus* or HIV* or AIDS or Acquired Immunodeficiency syndrome*).ti,ab. 599389  5 or/1-4 687072  6 (MSM or "men who have sex with men" or gay* or bisexual* or homosexual* or non-heterosexual*).ti,ab. 55286  7 homosexuality/ 18269  8 bisexuality/ 8821  9 men who have sex with men/ 16991  10 or/6-9 66330  11 circumcision/ 9938  12 prepuce/su [Surgery] 29  13 circumcis*.ti,ab. 9289  14 or/11-13 11824  15 5 and 10 and 14 301  16 limit 15 to yr="2018 - 2024" 80 | 10/16/24 | 80  -10  duplicates  = 70  unique items |
| **PsycINFO**  **(OVID)**  **1806-** | APA PsycInfo <1806 to October 2024 Week 2>  1 Human immunodeficiency virus/ 42567  2 acquired immune deficiency syndrome/ 16021  3 (Human immunodeficiency virus* or HIV* or AIDS or Acquired Immunodeficiency syndrome*).ti,ab. 79622  4 or/1-3 80427  5 (MSM or "men who have sex with men" or gay* or bisexual* or homosexual* or non-heterosexual*).ti,ab. 44935  6 homosexuality/ 8807  7 bisexuality/ 9557  8 men who have sex with men/ 4742  9 or/5-8 47670  10 circumcision/ 1025  11 circumcis*.ti,ab. 1177  12 or/10-11 1514  13 4 and 9 and 12 48  14 limit 13 to yr="2018 - 2024" 9 | 10/16/24 | 9  -1  duplicates  = 8  unique items |
| **Cochrane Library** | #1 MeSH descriptor: [HIV] explode all trees 4265  #2 MeSH descriptor: [HIV Infections] explode all trees 17997  #3 MeSH descriptor: [Acquired Immunodeficiency Syndrome] explode all trees 2498  #4 ("Human immunodeficiency virus" or HIV* or AIDS or "Acquired Immunodeficiency syndrome"):ti,ab 37750  #5 #1 OR #2 OR #3 OR #4 38888  #6 MeSH descriptor: [Homosexuality] explode all trees 943  #7 MeSH descriptor: [Bisexuality] explode all trees 96  #8 (MSM or "men who have sex with men" or gay* or bisexual* or homosexual* or "nonheterosexual"):ti,ab 2632  #9 #6 OR #7 OR #8 2746  #10 MeSH descriptor: [Circumcision, Male] explode all trees 415  #11 (circumcis*):ti,ab 936  #12 #10 OR #11 957  #13 #5 AND #9 AND #12 with Cochrane Library publication date Between Jan 2018 and Oct 2024 14 | 10/16/24 | 14  -6  duplicates  = 8  unique items |
| **Global Health**  **(OVID)**  **1910-** | Global Health <1910 to 2024 Week 42>  1 human immunodeficiency viruses/ 185436  2 acquired immune deficiency syndrome/ 62256  3 (Human immunodeficiency virus* or HIV* or AIDS or Acquired Immunodeficiency syndrome*).ti,ab. 227668  4 1 or 2 or 3 235783  5 (MSM or "men who have sex with men" or gay* or bisexual* or homosexual* or non-heterosexual*).ti,ab. 22604  6 homosexuality/ 14947  7 bisexuality/ 2331  8 5 or 6 or 7 24232  9 circumcision/ 1940  10 circumcis*.ti,ab. 2370  11 9 or 10 2641  12 4 and 8 and 11 109  13 limit 12 to yr="2018 - 2024" 28 | 10/16/24 | 28  -24  duplicates  = 4  unique items |
| **Scopus** | ( TITLE-ABS ( "Human immunodeficiency virus" OR hiv* OR "Acquired Immune deficiency virus" OR "Acquired immunodeficiency Virus" OR aids* ) ) AND ( TITLE-ABS ( "men who have sex with men" OR msm OR gay* OR homosexual* OR bisexual* OR "non-heterosexual" ) ) AND ( TITLE-ABS ( circumcis* ) ) AND PUBYEAR > 2017 AND PUBYEAR < 2025 | 10/16/24 | 37  -34  duplicates  = 3  unique items |

Notes: Duplicates were identified using the Endnote automated "find duplicates" function with preference set to match on title, author and year, and removed from your Endnote library. There will likely be additional duplicates found that Endnote was unable to detect.
