## Appendices for "Circumcision for HIV prevention in men who have sex with men: an updated meta-analysis": appendix 2 meta-analysis-final code.docx

VMMC Among Insertive Men

2025-05-27

#### Introduction

This document conducts a meta-analysis using the metafor package of 12 studies of VMMC among predominately insertive men. The data are processed to calculate log odds ratios (logOR) and their variances before performing a random-effects meta-analysis. Forest and funnel plots are generated to visualize the results, and bias is assessed using Egger’s test and the trim-and-fill method. Note that RR’s are converted to log ORs.

#### Load and Preprocess Data

The data are loaded from a CSV file, adjusted for missing or zero values, and filtered to exclude specific studies. Note that here I’m excluding Guanira because it’s unconfirmed.

```
# Load the data
meta_data <- read_csv("metaregression csv1.csv")
New names:
Rows: 32 Columns: 24
── Column specification
──────────────────────────────────────────────────────── Delimiter: "," chr
(4): Study, Group, Metaregression_Group, Region dbl (14): a, b, c, d, OR,
OR_LowerCI, OR_UpperCI, OR_p, RR, RR_LowerCI, RR_U... lgl (6): PR, PR_LowerCI,
PR_UpperCI, Insertive_Only, ...22, ...23
ℹ Use `spec()` to retrieve the full column specification for this data. ℹ
Specify the column types or set `show_col_types = FALSE` to quiet this message.
• `` -> `...22`
• `` -> `...23`
# Replace zeroes and filter data
meta_data <- meta_data %>%
  mutate(across(OR:PR_UpperCI, ~ if_else(. == 0, 0.01, .)),
    adjusted_measure = if_else(if_all(OR:PR_UpperCI, ~ is.na(.)), "Counts", "Adjusted")
  ) %>%
  filter(Metaregression_Group != "Exclude"  & !str_detect(Study, "Guanira"))

meta_data$region_group <- ifelse(
  meta_data$Region %in% c("Australia", "Europe","US/Canada"),
  "Aus_US_Can_Eur",
  "Other"
)

meta_data %>% gt::gt(.)
```

| Study | Group | a | b | c | d | OR | OR_LowerCI | OR_UpperCI | OR_p | RR | RR_LowerCI | RR_UpperCI | HR | HR_LowerCI | HR_UpperCI | PR | PR_LowerCI | PR_UpperCI | Insertive_Only | Metaregression_Group | ...22 | ...23 | Region | adjusted_measure | region_group |
| --- | --- | --- | --- | --- | --- | --- | --- | --- | --- | --- | --- | --- | --- | --- | --- | --- | --- | --- | --- | --- | --- | --- | --- | --- | --- |
| Reisen | Insertive | 2 | 14 | 28 | 48 | NA | NA | NA | NA | NA | NA | NA | NA | NA | NA | NA | NA | NA | TRUE | Ins | NA | NA | US/Canada | Counts | Aus_US_Can_Eur |
| Reisen | Other | 26 | 87 | 55 | 167 | NA | NA | NA | NA | NA | NA | NA | NA | NA | NA | NA | NA | NA | FALSE | Oth | NA | NA | US/Canada | Counts | Aus_US_Can_Eur |
| McDaid | Insertive | 0 | 2 | 27 | 77 | NA | NA | NA | NA | NA | NA | NA | NA | NA | NA | NA | NA | NA | TRUE | Ins | NA | NA | Europe | Counts | Aus_US_Can_Eur |
| McDaid | Everyone else | 2 | 17 | 55 | 296 | NA | NA | NA | NA | NA | NA | NA | NA | NA | NA | NA | NA | NA | FALSE | Oth | NA | NA | Europe | Counts | Aus_US_Can_Eur |
| Pando | Receptive | 7 | 37 | 24 | 117 | NA | NA | NA | NA | NA | NA | NA | NA | NA | NA | NA | NA | NA | FALSE | Oth | NA | NA | Latin America | Counts | Other |
| Pando | Insertive | 0 | 34 | 33 | 197 | NA | NA | NA | NA | NA | NA | NA | NA | NA | NA | NA | NA | NA | TRUE | Ins | NA | NA | Latin America | Counts | Other |
| Templeton | Insertive | 2 | 5 | 277 | 151 | NA | NA | NA | NA | NA | NA | NA | 0.11 | 0.01 | 0.92 | NA | NA | NA | TRUE | Ins | NA | NA | Australia | Adjusted | Aus_US_Can_Eur |
| Templeton | Receptive | 27 | 12 | 632 | 320 | NA | NA | NA | NA | NA | NA | NA | NA | NA | NA | NA | NA | NA | FALSE | Oth | NA | NA | Australia | Counts | Aus_US_Can_Eur |
| Sanchez | >=60% Insertive | NA | NA | NA | NA | NA | NA | NA | NA | 0.31 | 0.06 | 1.51 | NA | NA | NA | NA | NA | NA | TRUE | Ins | NA | NA | 25% US, 75% Latin America | Adjusted | Other |
| Qian | Everyone else | 9 | 177 | 38 | 364 | 0.54 | 0.25 | 1.14 | NA | NA | NA | NA | NA | NA | NA | NA | NA | NA | FALSE | Oth | NA | NA | Asia | Adjusted | Other |
| Qian | Predominant Insertive | 2 | 76 | 29 | 358 | 0.38 | 0.09 | 1.64 | NA | NA | NA | NA | NA | NA | NA | NA | NA | NA | TRUE | Ins | NA | NA | Asia | Adjusted | Other |
| Calzavara | Exclusive insertive | 7 | 6 | 17 | 8 | NA | NA | NA | NA | NA | NA | NA | NA | NA | NA | NA | NA | NA | TRUE | Ins | NA | NA | US/Canada | Counts | Aus_US_Can_Eur |
| Calzavara | Everyone else | 50 | 16 | 88 | 43 | NA | NA | NA | NA | NA | NA | NA | NA | NA | NA | NA | NA | NA | FALSE | Oth | NA | NA | US/Canada | Counts | Aus_US_Can_Eur |
| Doerner | Most and exclusively insertive - combined | 17 | 80 | 180 | 820 | 0.79 | 0.41 | 1.44 | NA | NA | NA | NA | NA | NA | NA | NA | NA | NA | TRUE | Ins | NA | NA | Europe | Adjusted | Aus_US_Can_Eur |
| Zeng | Exclusively and Predominantly Insertive | 6 | 26 | 32 | 246 | NA | NA | NA | NA | NA | NA | NA | NA | NA | NA | NA | NA | NA | TRUE | Ins | NA | NA | Asia | Counts | Other |
| Zeng | Everyone else | 3 | 39 | 19 | 149 | NA | NA | NA | NA | NA | NA | NA | NA | NA | NA | NA | NA | NA | FALSE | Oth | NA | NA | Asia | Counts | Other |
| Jameson | Only receptive | 115 | 17 | 326 | 83 | 1.47 | 0.94 | 2.33 | NA | NA | NA | NA | NA | NA | NA | NA | NA | NA | FALSE | Oth | NA | NA | US/Canada | Adjusted | Aus_US_Can_Eur |
| Jameson | Verse | 697 | 124 | 2240 | 434 | 1.08 | 0.98 | 1.18 | NA | NA | NA | NA | NA | NA | NA | NA | NA | NA | FALSE | Oth | NA | NA | US/Canada | Adjusted | Aus_US_Can_Eur |
| Jameson | Only insertive | 145 | 25 | 778 | 140 | 1.14 | 0.59 | 2.17 | NA | NA | NA | NA | NA | NA | NA | NA | NA | NA | TRUE | Ins | NA | NA | US/Canada | Adjusted | Aus_US_Can_Eur |
| Gao | Insertive | 0 | 5 | 124 | 118 | NA | NA | NA | NA | NA | NA | NA | 0.09 | 0.01 | 0.81 | NA | NA | NA | TRUE | Ins | NA | NA | Asia | Adjusted | Other |
| de Leon | Insertive | NA | NA | NA | NA | 0.29 | 0.19 | 0.44 | NA | NA | NA | NA | NA | NA | NA | NA | NA | NA | FALSE | Ins | NA | NA | Africa | Adjusted | Other |
| de Leon | Verse | NA | NA | NA | NA | 0.59 | 0.34 | 1.02 | NA | NA | NA | NA | NA | NA | NA | NA | NA | NA | FALSE | Oth | NA | NA | Africa | Adjusted | Other |
| de Leon | Receptive | NA | NA | NA | NA | 0.67 | 0.36 | 1.28 | NA | NA | NA | NA | NA | NA | NA | NA | NA | NA | FALSE | Oth | NA | NA | Africa | Adjusted | Other |
| Schneider | Insertive | 1 | 5 | 40 | 33 | NA | NA | NA | NA | NA | NA | NA | NA | NA | NA | NA | NA | NA | TRUE | Ins | NA | NA | Asia | Counts | Other |
| Schneider | Receptive | 16 | 44 | 62 | 114 | NA | NA | NA | NA | NA | NA | NA | NA | NA | NA | NA | NA | NA | FALSE | Oth | NA | NA | Asia | Counts | Other |
| Millett | Insertive (Black) | NA | NA | NA | NA | 1.47 | NA | NA | 0.25 | NA | NA | NA | NA | NA | NA | NA | NA | NA | TRUE | Ins | NA | NA | US/Canada | Adjusted | Aus_US_Can_Eur |
| Millett | Insertive (Latino) | NA | NA | NA | NA | 1.59 | NA | NA | 0.28 | NA | NA | NA | NA | NA | NA | NA | NA | NA | TRUE | Ins | NA | NA | US/Canada | Adjusted | Aus_US_Can_Eur |

#### Calculate Log Odds Ratios and Variances

We calculate the log odds ratios (logOR) and variances in three steps: using contingency tables, reported ORs, and approximations for RR, HR, and PR.

##### Step 1: Using Contingency Tables

```
meta_data <- meta_data %>%
  mutate(logOR_cell_count = NA, varOR_cell_count = NA)

for (i in 1:nrow(meta_data)) {
  if (!is.na(meta_data$a[i]) && !is.na(meta_data$b[i]) && !is.na(meta_data$c[i]) && !is.na(meta_data$d[i])) {
    result <- escalc(
      measure = "OR",
      ai = meta_data$a[i], bi = meta_data$b[i],
      ci = meta_data$c[i], di = meta_data$d[i]
    )
    meta_data$logOR_cell_count[i] <- result$yi
    meta_data$varOR_cell_count[i] <- result$vi
  }
}
```

##### Step 2: Using Reported ORs

```
meta_data <- meta_data %>%
  mutate(logOR_effect_measure = NA, varOR_effect_measure = NA)

for (i in seq_len(nrow(meta_data))) {
  if (!is.na(meta_data$OR[i])) {
    this_logOR <- log(meta_data$OR[i])
    meta_data$logOR_effect_measure[i] <- this_logOR

    if (!is.na(meta_data$OR_LowerCI[i]) && !is.na(meta_data$OR_UpperCI[i])) {
      logCI_lower <- log(meta_data$OR_LowerCI[i])
      logCI_upper <- log(meta_data$OR_UpperCI[i])
      se_logOR <- (logCI_upper - logCI_lower) / (2 * 1.96)
      meta_data$varOR_effect_measure[i] <- se_logOR^2
    } else if (!is.na(meta_data$OR_p[i])) {
      p_val <- meta_data$OR_p[i]
      z_val <- qnorm(1 - p_val / 2)
      se_logOR <- abs(this_logOR) / z_val
      meta_data$varOR_effect_measure[i] <- se_logOR^2
    }
  }
}
```

##### Step 3: Converting RRs, HRs, and PRs

```
for (i in 1:nrow(meta_data)) {
  if (!is.na(meta_data$RR[i])) {
    meta_data$logOR_effect_measure[i] <- log(meta_data$RR[i])
    meta_data$varOR_effect_measure[i] <- ((log(meta_data$RR_UpperCI[i]) - log(meta_data$RR_LowerCI[i])) / (2 * 1.96))^2
  }
  if (!is.na(meta_data$HR[i])) {
    meta_data$logOR_effect_measure[i] <- log(meta_data$HR[i])
    meta_data$varOR_effect_measure[i] <- ((log(meta_data$HR_UpperCI[i]) - log(meta_data$HR_LowerCI[i])) / (2 * 1.96))^2
  }
  if (!is.na(meta_data$PR[i])) {
    meta_data$logOR_effect_measure[i] <- log(meta_data$PR[i])
    meta_data$varOR_effect_measure[i] <- ((log(meta_data$PR_UpperCI[i]) - log(meta_data$PR_LowerCI[i])) / (2 * 1.96))^2
  }
}
meta_data <- meta_data %>%
  mutate(
    logOR_prefer_cell_count = if_else(!is.na(logOR_cell_count), logOR_cell_count, logOR_effect_measure),
    varOR_prefer_cell_count = if_else(!is.na(varOR_cell_count), varOR_cell_count, varOR_effect_measure),
    logOR_prefer_adjusted = if_else(!is.na(logOR_effect_measure), logOR_effect_measure, logOR_cell_count),
    varOR_prefer_adjusted = if_else(!is.na(varOR_effect_measure), varOR_effect_measure, varOR_cell_count)
  )
```

#### Perform Random-Effects Meta-Analysis for Insertive Only

This includes de Leon, but not excluded later.

```
# Filter data to include only insertive groups
meta_data_insert_only <- meta_data %>% filter(Insertive_Only)

# Analysis using cell counts only
meta_results_cell_count <- rma.mv(
  yi = logOR_cell_count,
  V = varOR_cell_count,
  data = meta_data_insert_only,
  method = "REML",
  random = ~ 1 | Study
)
Warning: 3 rows with NAs omitted from model fitting.
# Analysis using cell counts and effect measures, preferring cell counts
meta_results_prefer_cell_count <- rma.mv(
  yi = logOR_prefer_cell_count,
  V = varOR_prefer_cell_count,
  data = meta_data_insert_only,
  method = "REML",
  random = ~ 1 | Study
)

# Analysis using cell counts and effect measures, preferring adjusted effect measures
meta_results_prefer_adjusted <- rma.mv(
  yi = logOR_prefer_adjusted,
  V = varOR_prefer_adjusted,
  data = meta_data_insert_only,
  method = "REML",
  random = ~ 1 | Study
)

# Transform results back to OR scale
pooled_or_cell_count <- exp(meta_results_cell_count$b)
lower_ci_cell_count <- exp(meta_results_cell_count$ci.lb)
upper_ci_cell_count <- exp(meta_results_cell_count$ci.ub)

pooled_or_prefer_cell_count <- exp(meta_results_prefer_cell_count$b)
lower_ci_prefer_cell_count <- exp(meta_results_prefer_cell_count$ci.lb)
upper_ci_prefer_cell_count <- exp(meta_results_prefer_cell_count$ci.ub)

pooled_or_prefer_adjusted <- exp(meta_results_prefer_adjusted$b)
lower_ci_prefer_adjusted <- exp(meta_results_prefer_adjusted$ci.lb)
upper_ci_prefer_adjusted <- exp(meta_results_prefer_adjusted$ci.ub)

# Display results
pooled_or_results <- data.frame(
  Analysis = c("Cell Count Only", "Prefer Cell Count", "Prefer Adjusted"),
  Pooled_OR = round(c(pooled_or_cell_count, pooled_or_prefer_cell_count, pooled_or_prefer_adjusted), 3),
  Lower_95_CI = round(c(lower_ci_cell_count, lower_ci_prefer_cell_count, lower_ci_prefer_adjusted), 3),
  Upper_95_CI = round(c(upper_ci_cell_count, upper_ci_prefer_cell_count, upper_ci_prefer_adjusted), 3)
)

pooled_or_results %>% gt::gt(.)
```

| Analysis | Pooled_OR | Lower_95_CI | Upper_95_CI |
| --- | --- | --- | --- |
| Cell Count Only | 0.56 | 0.319 | 0.983 |
| Prefer Cell Count | 0.62 | 0.374 | 1.029 |
| Prefer Adjusted | 0.57 | 0.331 | 0.981 |

```
# Sort the data by effect size (log odds ratio)
meta_data_sorted <- meta_data_insert_only %>%
  arrange(logOR_prefer_adjusted)

# Perform Random-Effects Meta-Analysis using the sorted data
meta_results_prefer_adjusted_sorted <- rma.mv(
  yi = logOR_prefer_adjusted,
  V = varOR_prefer_adjusted,
  data = meta_data_sorted,
  method = "REML",
  random = ~ 1 | Study
)
meta_results_prefer_adjusted_sorted

Multivariate Meta-Analysis Model (k = 14; method: REML)

Variance Components:

            estim    sqrt  nlvls  fixed  factor
sigma^2    0.4667  0.6832     13     no   Study

Test for Heterogeneity:
Q(df = 13) = 25.2963, p-val = 0.0211

Model Results:

estimate      se     zval    pval    ci.lb    ci.ub
 -0.5622  0.2770  -2.0301  0.0423  -1.1050  -0.0194  *

---
Signif. codes:  0 '***' 0.001 '**' 0.01 '*' 0.05 '.' 0.1 ' ' 1
```

#### Visualize Results

##### Forest Plot

```
# Create a forest plot sorted by effect size
forest(meta_results_prefer_adjusted_sorted,
  atransf = exp, slab = meta_data_sorted$Study,
  xlab = "Odds Ratio (OR)", refline = 0
)
```

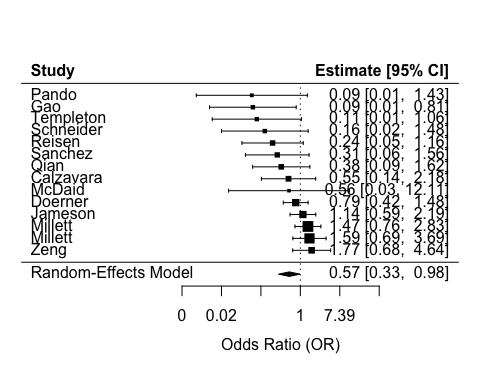

##### Funnel Plot

funnel(meta_results_prefer_adjusted, xlab = "Effect Size (Log Odds Ratio)", ylab = "Standard Error")

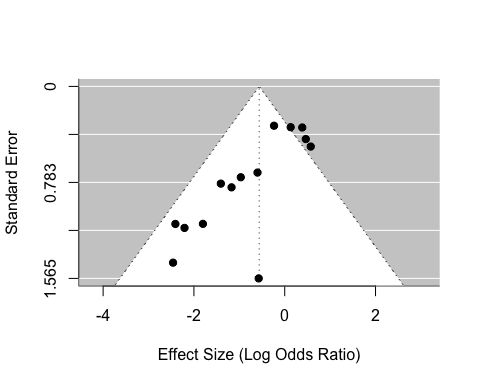

#### **Subgroup Analysis**

We filter the data to include only rows from studies that have both insertive and other groups. We also set “Oth” as the reference for the Metaregression_Group.

```
# Load necessary libraries
library(metafor)
library(tidyverse)

# Filter data to include only studies with both insertive and other groups
meta_data_reg <- meta_data %>%
  mutate(
    # Make 'Oth' the reference for Metaregression_Group
    Metaregression_Group = relevel(factor(Metaregression_Group), ref = "Oth")
  ) %>%
  group_by(Study) %>%
  filter(n() > 1 & !str_detect(Study, "Millet")) %>%
  ungroup()
```

For clarity, the included data from 10 studiescan be seen below

meta_data_reg %>% gt::gt(.)

| Study | Group | a | b | c | d | OR | OR_LowerCI | OR_UpperCI | OR_p | RR | RR_LowerCI | RR_UpperCI | HR | HR_LowerCI | HR_UpperCI | PR | PR_LowerCI | PR_UpperCI | Insertive_Only | Metaregression_Group | ...22 | ...23 | Region | adjusted_measure | region_group | logOR_cell_count | varOR_cell_count | logOR_effect_measure | varOR_effect_measure | logOR_prefer_cell_count | varOR_prefer_cell_count | logOR_prefer_adjusted | varOR_prefer_adjusted |
| --- | --- | --- | --- | --- | --- | --- | --- | --- | --- | --- | --- | --- | --- | --- | --- | --- | --- | --- | --- | --- | --- | --- | --- | --- | --- | --- | --- | --- | --- | --- | --- | --- | --- |
| Reisen | Insertive | 2 | 14 | 28 | 48 | NA | NA | NA | NA | NA | NA | NA | NA | NA | NA | NA | NA | NA | TRUE | Ins | NA | NA | US/Canada | Counts | Aus_US_Can_Eur | -1.40691365 | 0.62797619 | NA | NA | -1.40691365 | 0.62797619 | -1.40691365 | 0.627976190 |
| Reisen | Other | 26 | 87 | 55 | 167 | NA | NA | NA | NA | NA | NA | NA | NA | NA | NA | NA | NA | NA | FALSE | Oth | NA | NA | US/Canada | Counts | Aus_US_Can_Eur | -0.09715095 | 0.07412563 | NA | NA | -0.09715095 | 0.07412563 | -0.09715095 | 0.074125633 |
| McDaid | Insertive | 0 | 2 | 27 | 77 | NA | NA | NA | NA | NA | NA | NA | NA | NA | NA | NA | NA | NA | TRUE | Ins | NA | NA | Europe | Counts | Aus_US_Can_Eur | -0.57334598 | 2.44926686 | NA | NA | -0.57334598 | 2.44926686 | -0.57334598 | 2.449266862 |
| McDaid | Everyone else | 2 | 17 | 55 | 296 | NA | NA | NA | NA | NA | NA | NA | NA | NA | NA | NA | NA | NA | FALSE | Oth | NA | NA | Europe | Counts | Aus_US_Can_Eur | -0.45703989 | 0.58038373 | NA | NA | -0.45703989 | 0.58038373 | -0.45703989 | 0.580383726 |
| Pando | Receptive | 7 | 37 | 24 | 117 | NA | NA | NA | NA | NA | NA | NA | NA | NA | NA | NA | NA | NA | FALSE | Oth | NA | NA | Latin America | Counts | Other | -0.08088766 | 0.22009785 | NA | NA | -0.08088766 | 0.22009785 | -0.08088766 | 0.220097845 |
| Pando | Insertive | 0 | 34 | 33 | 197 | NA | NA | NA | NA | NA | NA | NA | NA | NA | NA | NA | NA | NA | TRUE | Ins | NA | NA | Latin America | Counts | Other | -2.45991336 | 2.06389954 | NA | NA | -2.45991336 | 2.06389954 | -2.45991336 | 2.063899545 |
| Templeton | Insertive | 2 | 5 | 277 | 151 | NA | NA | NA | NA | NA | NA | NA | 0.11 | 0.01 | 0.92 | NA | NA | NA | TRUE | Ins | NA | NA | Australia | Adjusted | Aus_US_Can_Eur | -1.52302840 | 0.71023262 | -2.20727491 | 1.330602609 | -1.52302840 | 0.71023262 | -2.20727491 | 1.330602609 |
| Templeton | Receptive | 27 | 12 | 632 | 320 | NA | NA | NA | NA | NA | NA | NA | NA | NA | NA | NA | NA | NA | FALSE | Oth | NA | NA | Australia | Counts | Aus_US_Can_Eur | 0.13036182 | 0.12507765 | NA | NA | 0.13036182 | 0.12507765 | 0.13036182 | 0.125077649 |
| Qian | Everyone else | 9 | 177 | 38 | 364 | 0.54 | 0.25 | 1.14 | NA | NA | NA | NA | NA | NA | NA | NA | NA | NA | FALSE | Oth | NA | NA | Asia | Adjusted | Other | -0.71935745 | 0.14582387 | -0.61618614 | 0.149824809 | -0.71935745 | 0.14582387 | -0.61618614 | 0.149824809 |
| Qian | Predominant Insertive | 2 | 76 | 29 | 358 | 0.38 | 0.09 | 1.64 | NA | NA | NA | NA | NA | NA | NA | NA | NA | NA | TRUE | Ins | NA | NA | Asia | Adjusted | Other | -1.12434900 | 0.55043395 | -0.96758403 | 0.548295613 | -1.12434900 | 0.55043395 | -0.96758403 | 0.548295613 |
| Calzavara | Exclusive insertive | 7 | 6 | 17 | 8 | NA | NA | NA | NA | NA | NA | NA | NA | NA | NA | NA | NA | NA | TRUE | Ins | NA | NA | US/Canada | Counts | Aus_US_Can_Eur | -0.59962112 | 0.49334734 | NA | NA | -0.59962112 | 0.49334734 | -0.59962112 | 0.493347339 |
| Calzavara | Everyone else | 50 | 16 | 88 | 43 | NA | NA | NA | NA | NA | NA | NA | NA | NA | NA | NA | NA | NA | FALSE | Oth | NA | NA | US/Canada | Counts | Aus_US_Can_Eur | 0.42329758 | 0.11711945 | NA | NA | 0.42329758 | 0.11711945 | 0.42329758 | 0.117119450 |
| Zeng | Exclusively and Predominantly Insertive | 6 | 26 | 32 | 246 | NA | NA | NA | NA | NA | NA | NA | NA | NA | NA | NA | NA | NA | TRUE | Ins | NA | NA | Asia | Counts | Other | 0.57325856 | 0.24044325 | NA | NA | 0.57325856 | 0.24044325 | 0.57325856 | 0.240443246 |
| Zeng | Everyone else | 3 | 39 | 19 | 149 | NA | NA | NA | NA | NA | NA | NA | NA | NA | NA | NA | NA | NA | FALSE | Oth | NA | NA | Asia | Counts | Other | -0.50544203 | 0.41831735 | NA | NA | -0.50544203 | 0.41831735 | -0.50544203 | 0.418317347 |
| Jameson | Only receptive | 115 | 17 | 326 | 83 | 1.47 | 0.94 | 2.33 | NA | NA | NA | NA | NA | NA | NA | NA | NA | NA | FALSE | Oth | NA | NA | US/Canada | Adjusted | Aus_US_Can_Eur | 0.54366201 | 0.08263486 | 0.38526240 | 0.053623397 | 0.54366201 | 0.08263486 | 0.38526240 | 0.053623397 |
| Jameson | Verse | 697 | 124 | 2240 | 434 | 1.08 | 0.98 | 1.18 | NA | NA | NA | NA | NA | NA | NA | NA | NA | NA | FALSE | Oth | NA | NA | US/Canada | Adjusted | Aus_US_Can_Eur | 0.08531723 | 0.01224981 | 0.07696104 | 0.002244563 | 0.08531723 | 0.01224981 | 0.07696104 | 0.002244563 |
| Jameson | Only insertive | 145 | 25 | 778 | 140 | 1.14 | 0.59 | 2.17 | NA | NA | NA | NA | NA | NA | NA | NA | NA | NA | TRUE | Ins | NA | NA | US/Canada | Adjusted | Aus_US_Can_Eur | 0.04277382 | 0.05532476 | 0.13102826 | 0.110379876 | 0.04277382 | 0.05532476 | 0.13102826 | 0.110379876 |
| de Leon | Insertive | NA | NA | NA | NA | 0.29 | 0.19 | 0.44 | NA | NA | NA | NA | NA | NA | NA | NA | NA | NA | FALSE | Ins | NA | NA | Africa | Adjusted | Other | NA | NA | -1.23787436 | 0.045891111 | -1.23787436 | 0.04589111 | -1.23787436 | 0.045891111 |
| de Leon | Verse | NA | NA | NA | NA | 0.59 | 0.34 | 1.02 | NA | NA | NA | NA | NA | NA | NA | NA | NA | NA | FALSE | Oth | NA | NA | Africa | Adjusted | Other | NA | NA | -0.52763274 | 0.078544679 | -0.52763274 | 0.07854468 | -0.52763274 | 0.078544679 |
| de Leon | Receptive | NA | NA | NA | NA | 0.67 | 0.36 | 1.28 | NA | NA | NA | NA | NA | NA | NA | NA | NA | NA | FALSE | Oth | NA | NA | Africa | Adjusted | Other | NA | NA | -0.40047757 | 0.104716849 | -0.40047757 | 0.10471685 | -0.40047757 | 0.104716849 |
| Schneider | Insertive | 1 | 5 | 40 | 33 | NA | NA | NA | NA | NA | NA | NA | NA | NA | NA | NA | NA | NA | TRUE | Ins | NA | NA | Asia | Counts | Other | -1.80180981 | 1.25530303 | NA | NA | -1.80180981 | 1.25530303 | -1.80180981 | 1.255303030 |
| Schneider | Receptive | 16 | 44 | 62 | 114 | NA | NA | NA | NA | NA | NA | NA | NA | NA | NA | NA | NA | NA | FALSE | Oth | NA | NA | Asia | Counts | Other | -0.40253685 | 0.11012823 | NA | NA | -0.40253685 | 0.11012823 | -0.40253685 | 0.110128235 |

We perform subgroup analyses for insertive and other groups separately and display the pooled odds ratios and confidence intervals for each subgroup.

```
# Split data into two subgroups
ins_data <- meta_data_reg %>% filter(Metaregression_Group == "Ins")
oth_data <- meta_data_reg %>% filter(Metaregression_Group == "Oth")

# Subgroup meta-analyses
res_ins <- rma(yi = logOR_prefer_adjusted, vi = varOR_prefer_adjusted, data = ins_data, method = "REML")
res_oth <- rma(yi = logOR_prefer_adjusted, vi = varOR_prefer_adjusted, data = oth_data, method = "REML")

# Show pooled OR + 95% CI for each subgroup
subgroup_or <- bind_rows(
  data.frame(
    Group    = "Other (Reference)",
    OR       = round(exp(res_oth$b[1]), 3),
    CI_lower = round(exp(res_oth$ci.lb[1]), 3),
    CI_upper = round(exp(res_oth$ci.ub[1]), 3)
  ),
  data.frame(
    Group    = "Insertive",
    OR       = round(exp(res_ins$b[1]), 3),
    CI_lower = round(exp(res_ins$ci.lb[1]), 3),
    CI_upper = round(exp(res_ins$ci.ub[1]), 3)
  )
)

cat("\nSubgroup-Specific Pooled ORs:\n")

Subgroup-Specific Pooled ORs:
print(subgroup_or)
              Group    OR CI_lower CI_upper
1 Other (Reference) 0.924    0.748    1.142
2         Insertive 0.460    0.246    0.860
```

### By Region

Here we order the Odds ratios by their variance within region and see trends toward decreasing effect as the variance gets smaller. This is indicative of publication bias.

```
meta_data_sorted2 <- meta_data_insert_only %>%
  arrange(region_group,-varOR_prefer_adjusted)

# Perform Random-Effects Meta-Analysis using the sorted data
meta_results_prefer_adjusted_sorted <- rma.mv(
  yi = logOR_prefer_adjusted,
  V = varOR_prefer_adjusted,
  data = meta_data_sorted2,
  method = "REML",
  random = ~ 1 | Study
)

# Create a forest plot sorted by effect size
forest(meta_results_prefer_adjusted_sorted,
  atransf = exp, slab = paste0(meta_data_sorted2$Study, " (",meta_data_sorted2$Region,")") ,
  xlab = "Odds Ratio (OR)", refline = 0
)
```

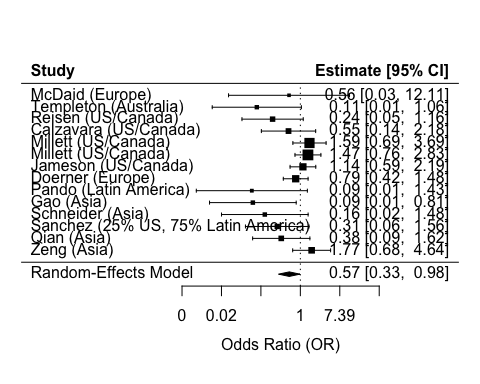

Here is the bias test for Aus-Eur-US-Can:

regtest(
x = logOR_prefer_adjusted,
vi = varOR_prefer_adjusted,
data = meta_data_sorted[meta_data_insert_only$region_group == "Aus_US_Can_Eur",])

Regression Test for Funnel Plot Asymmetry

Model: mixed-effects meta-regression model
Predictor: standard error

Test for Funnel Plot Asymmetry: z = -2.8338, p = 0.0046
Limit Estimate (as sei -> 0): b = 1.1803 (CI: 0.0295, 2.3310)

Here is the bias test for non-Aus-Eur-US-Can:

regtest(
x = logOR_prefer_adjusted,
vi = varOR_prefer_adjusted,
data = meta_data_sorted[meta_data_insert_only$region_group == "Other",])

Regression Test for Funnel Plot Asymmetry

Model: mixed-effects meta-regression model
Predictor: standard error

Test for Funnel Plot Asymmetry: z = -2.7425, p = 0.0061
Limit Estimate (as sei -> 0): b = 0.9453 (CI: 0.1177, 1.7730)

Here is the bias test for all combined:

regtest(
x = logOR_prefer_adjusted,
vi = varOR_prefer_adjusted,
data = meta_data_sorted)

Regression Test for Funnel Plot Asymmetry

Model: mixed-effects meta-regression model
Predictor: standard error

Test for Funnel Plot Asymmetry: z = -3.9798, p < .0001
Limit Estimate (as sei -> 0): b = 0.9708 (CI: 0.3445, 1.5972)

### Confounder Adjusted Analysis

Now we compare the odds ratio for hiv infection of insertive v.s. the study group with the lowest level of insertivity. For some studies this is non-insertive and for some it is a combination of non-insertive and verse.The comparison metric is the ratio of the odds ratios. We assume independence of the two odds ratios, which is true when it is constructed from counts and approximate otherwise.

Let $P^{i}\left( H|C,R \right)$ be the probability in study population $i$ of HIV positivity ($H\in\{0,1\}$) given circumcision status ($C\in\{0,1\}$) and the proportion of unprotected sexual contacts that are insertive ($R\in\left[ 0,1 \right]$). We model the odds logistically as

$$\log\left( \frac{P^{i}\left( H|C=c,R=r \right)}{1-P^{i}\left( H|C=c,R=r \right)} \right)=\beta_{0}^{i}+\beta_{1}^{i}c+g^{i}\left( r \right)+\beta_{2}^{i}f^{i}\left( r \right)c,$$

where $\beta_{0}^{i}$ models the baseline risk of HIV, and $g^{i}$ is an arbitrary function representing the effect of insertivity on HIV risk. $\beta_{1}^{i}$ models the protection of circumcision in purely non-insertive individuals. That is to say the social and personal effect of circumcision on HIV risk seperate from any real protection during insertive sex. We call this the confounding effect of circumcision.

$f^{i}$ in the last term is a monotonic increasing function from 0 to 1 ($f^{i}\left( 0 \right)=0$ and $f^{i}\left( 1 \right)=1$) mapping the relative protection given to people of mixed sexual roles. $\beta_{2}^{i}$ is then the “unconfounded” effect of circumcision on purely insertive individuals.

The first comparison we might do is to look at the log ratio of insertive circumcised v.s. insertive uncircumcised. In our model this would be represented as

$$\text{lr}^{i}\left( r \right)=\log\left( \frac{P^{i}\left( H|C=1,R=r \right)}{1-P^{i}\left( H|C=1,R=r \right)} \right)-\log\left( \frac{P^{i}\left( H|C=0,R=r \right)}{1-P^{i}\left( H|C=0,R=r \right)} \right)=\beta_{1}^{i}+\beta_{2}^{i}f^{i}\left( r \right)$$

We see that the log odds ratio includes the confounding term $\beta_{1}^{i}$ and the true effect term $\beta_{2}^{i}$ reduced by the level of non-insertivity ($f^{i}\left( r \right)$). Having both $\beta$s in the ratio means that we must assume no confounding effects ($\beta_{1}^{i}=0$) if we wish to interpret this ratio as the biological effect of circumcision.

In order to remove the confounding effect, we can look at the log ratio of high insertivity subtracting off the log ratio of a lower insertivity level

$$\text{lr}^{i}\left( r_{high} \right)-\text{lr}^{i}\left( r_{low} \right)=\beta_{2}^{i}\left( f^{i}\left( r_{high} \right)-f^{i}\left( r_{low} \right) \right).$$

This term includes only the real effect of circumcision $\beta_{2}^{i}$, removing the confounding effect $\beta_{1}^{i}$. The full magnitude of the $\beta_{2}^{i}$ effect does not translate to the log ratio of ratios, rather its magnitude is reduced by a factor of $0\leq\left( f^{i}\left( r_{high} \right)-f^{i}\left( r_{low} \right) \right)\leq1$.

For each study we calculate the difference of log odds ratios between the most and least insertive groups reported. The variance of this difference is calculated assuming independence, which is exactly true when the odds ratios are calculated from counts and approximately true otherwise.

```
meta_comp <- meta_data |>
  filter(Group != "Verse" & Study != "Millett") |>
  mutate(lor = logOR_prefer_adjusted, vl=varOR_prefer_adjusted) |>
  select(Study,Metaregression_Group,lor,vl, region_group) |>
  pivot_wider(
    names_from=Metaregression_Group,
    values_from=c(lor,vl))

meta_comp$com_lor <- meta_comp$lor_Ins - meta_comp$lor_Oth
meta_comp$com_vl <- meta_comp$vl_Ins + meta_comp$vl_Oth
meta_comp <- meta_comp |> arrange(-com_vl)
meta_results_comp <- rma.mv(
  yi = com_lor,
  V = com_vl,
  data = meta_comp,
  method = "REML",
  random = ~ 1 | Study
)
Warning: 3 rows with NAs omitted from model fitting.
meta_results_comp

Multivariate Meta-Analysis Model (k = 10; method: REML)

Variance Components:

            estim    sqrt  nlvls  fixed  factor
sigma^2    0.0274  0.1655     10     no   Study

Test for Heterogeneity:
Q(df = 9) = 10.4638, p-val = 0.3143

Model Results:

estimate      se     zval    pval    ci.lb    ci.ub
 -0.6389  0.2308  -2.7680  0.0056  -1.0914  -0.1865  **

---
Signif. codes:  0 '***' 0.001 '**' 0.01 '*' 0.05 '.' 0.1 ' ' 1
```

Note the high signficance of the result and large effect. Also note the lack of heterogeneity, which is good.

```
forest(meta_results_comp,
  atransf = exp, slab = paste0(meta_comp$Study) ,
  xlab = "Odds Ratio Ratio (Insertive OR / Other OR)", refline = 0
)
```

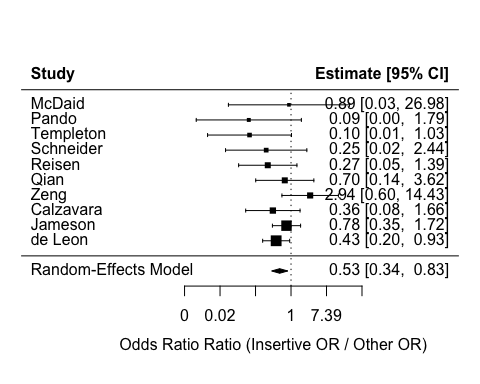

The forest plot is ordered by variance and shows no trends, which is confirmed by the funnel plot and bias test

```
funnel(
  meta_results_comp,
  xlab = "Log Odds Ratio - Log Odds Ratio",
  ylab = "Standard Error",
  main = "Funnel Plot: Insertive v.s. Other"
)
```

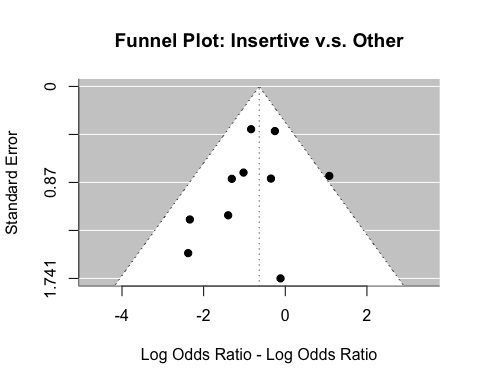

regtest(
x = com_lor,
vi = com_vl,
data = meta_comp)

Warning: 3 studies with NAs omitted from test.

Regression Test for Funnel Plot Asymmetry

Model: mixed-effects meta-regression model
Predictor: standard error

Test for Funnel Plot Asymmetry: z = -0.9640, p = 0.3351
Limit Estimate (as sei -> 0): b = -0.1598 (CI: -1.2920, 0.9725)

results are unchanged when removing de Leon:

```
#rma.mv(
#  yi = com_lor,
#  V = com_vl,
#  data = meta_comp[meta_comp$Study != "de Leon",],
#  #method = "REML",
#  random = ~ 1 | Study
#)

with(meta_comp[meta_comp$Study != "de Leon",], rma(
  yi = com_lor,
  vi = com_vl,
  #method = "REML",
  #random = ~ 1 | Study
))
Warning: 3 studies with NAs omitted from model fitting.

Random-Effects Model (k = 9; tau^2 estimator: REML)

tau^2 (estimated amount of total heterogeneity): 0.1870 (SE = 0.4322)
tau (square root of estimated tau^2 value):      0.4324
I^2 (total heterogeneity / total variability):   20.41%
H^2 (total variability / sampling variability):  1.26

Test for Heterogeneity:
Q(df = 8) = 10.0528, p-val = 0.2613

Model Results:

estimate      se     zval    pval    ci.lb   ci.ub
 -0.6231  0.3243  -1.9215  0.0547  -1.2587  0.0125  .

---
Signif. codes:  0 '***' 0.001 '**' 0.01 '*' 0.05 '.' 0.1 ' ' 1
```

### Regional Effect

```
meta_region_effect <- rma.mv(
  yi = logOR_prefer_adjusted,
  V = varOR_prefer_adjusted,
  data = meta_data_sorted2,
  method = "REML",
  mods = ~ region_group,
  random = ~ 1 | Study
)

summary(meta_region_effect)

Multivariate Meta-Analysis Model (k = 14; method: REML)

  logLik  Deviance       AIC       BIC      AICc
-16.4357   32.8713   38.8713   40.3261   41.8713

Variance Components:

            estim    sqrt  nlvls  fixed  factor
sigma^2    0.4765  0.6903     13     no   Study

Test for Residual Heterogeneity:
QE(df = 12) = 22.7373, p-val = 0.0300

Test of Moderators (coefficient 2):
QM(df = 1) = 1.1362, p-val = 0.2865

Model Results:

                   estimate      se     zval    pval    ci.lb   ci.ub
intrcpt             -0.3378  0.3515  -0.9609  0.3366  -1.0267  0.3512
region_groupOther   -0.6147  0.5766  -1.0659  0.2865  -1.7448  0.5155

---
Signif. codes:  0 '***' 0.001 '**' 0.01 '*' 0.05 '.' 0.1 ' ' 1
```

No statistically significant difference by region.

Running each region individually we get this for other:

```
meta_region_effect_oth <- rma.mv(
  yi = logOR_prefer_adjusted,
  V = varOR_prefer_adjusted,
  data = meta_data_sorted2[meta_data_sorted2$region_group == "Other",],
  method = "REML",
  random = ~ 1 | Study
)

summary(meta_region_effect_oth)

Multivariate Meta-Analysis Model (k = 6; method: REML)

  logLik  Deviance       AIC       BIC      AICc
 -8.1663   16.3325   20.3325   19.5514   26.3325

Variance Components:

            estim    sqrt  nlvls  fixed  factor
sigma^2    0.8845  0.9405      6     no   Study

Test for Heterogeneity:
Q(df = 5) = 11.8804, p-val = 0.0365

Model Results:

estimate      se     zval    pval    ci.lb    ci.ub
 -1.0617  0.5345  -1.9864  0.0470  -2.1092  -0.0141  *

---
Signif. codes:  0 '***' 0.001 '**' 0.01 '*' 0.05 '.' 0.1 ' ' 1
```

And this for Aus_US_Can_Eur

```
meta_region_effect_eur <- rma.mv(
  yi = logOR_prefer_adjusted,
  V = varOR_prefer_adjusted,
  data = meta_data_sorted2[meta_data_sorted2$region_group != "Other",],
  method = "REML",
  random = ~ 1 | Study
)

summary(meta_region_effect_eur)

Multivariate Meta-Analysis Model (k = 8; method: REML)

  logLik  Deviance       AIC       BIC      AICc
 -7.9103   15.8205   19.8205   19.7124   22.8205

Variance Components:

            estim    sqrt  nlvls  fixed  factor
sigma^2    0.1868  0.4322      7     no   Study

Test for Heterogeneity:
Q(df = 7) = 10.8568, p-val = 0.1450

Model Results:

estimate      se     zval    pval    ci.lb   ci.ub
 -0.2101  0.2630  -0.7986  0.4245  -0.7256  0.3055

---
Signif. codes:  0 '***' 0.001 '**' 0.01 '*' 0.05 '.' 0.1 ' ' 1
```

#### Regional effect: Confounder Adjusted

```
meta_results_comp_reg <- rma.mv(
  yi = com_lor,
  V = com_vl,
  data = meta_comp,
  method = "REML",
  mod = ~ region_group,
  random = ~ 1 | Study
)
Warning: 3 rows with NAs omitted from model fitting.
meta_results_comp

Multivariate Meta-Analysis Model (k = 10; method: REML)

Variance Components:

            estim    sqrt  nlvls  fixed  factor
sigma^2    0.0274  0.1655     10     no   Study

Test for Heterogeneity:
Q(df = 9) = 10.4638, p-val = 0.3143

Model Results:

estimate      se     zval    pval    ci.lb    ci.ub
 -0.6389  0.2308  -2.7680  0.0056  -1.0914  -0.1865  **

---
Signif. codes:  0 '***' 0.001 '**' 0.01 '*' 0.05 '.' 0.1 ' ' 1
```

Confounder adjusted effect in non-Aus_US_Can_Eur regions

```
rma.mv(
  yi = com_lor,
  V = com_vl,
  data = meta_comp[meta_comp$region_group == "Other",],
  method = "REML",
  random = ~ 1 | Study
)
Warning: 2 rows with NAs omitted from model fitting.

Multivariate Meta-Analysis Model (k = 5; method: REML)

Variance Components:

            estim    sqrt  nlvls  fixed  factor
sigma^2    0.3796  0.6161      5     no   Study

Test for Heterogeneity:
Q(df = 4) = 6.5940, p-val = 0.1590

Model Results:

estimate      se     zval    pval    ci.lb   ci.ub
 -0.5421  0.4600  -1.1784  0.2386  -1.4437  0.3595

---
Signif. codes:  0 '***' 0.001 '**' 0.01 '*' 0.05 '.' 0.1 ' ' 1
```

Confounder adjusted effect in Aus_US_Can_Eur regions

```
rma.mv(
  yi = com_lor,
  V = com_vl,
  data = meta_comp[meta_comp$region_group != "Other",],
  method = "REML",
  random = ~ 1 | Study
)
Warning: 1 row with NAs omitted from model fitting.

Multivariate Meta-Analysis Model (k = 5; method: REML)

Variance Components:

            estim    sqrt  nlvls  fixed  factor
sigma^2    0.1280  0.3578      5     no   Study

Test for Heterogeneity:
Q(df = 4) = 3.8520, p-val = 0.4264

Model Results:

estimate      se     zval    pval    ci.lb    ci.ub
 -0.7714  0.3798  -2.0308  0.0423  -1.5158  -0.0269  *

---
Signif. codes:  0 '***' 0.001 '**' 0.01 '*' 0.05 '.' 0.1 ' ' 1
```
