## Appendices for "Circumcision for HIV prevention in men who have sex with men: an updated meta-analysis": appendix 4 additional results.docx

**Online appendix 4: Supplementary Results – included data and descriptive analysis**

*Included studies:* Sixteen of 49 studies were not included the Yuan et al meta-analysis; 21 were not included in Zheng et al.

*Descriptive analysis:* Two included studies stratified by whether participants also had sex with women^48,50,^ Seventeen provided analyses adjusted for at least one measure of risk behavior, and six adjusted for STI outcomes^26,35,37,44,54,63^. Three relied on self-report for assessing HIV status^30,43,54^; for circumcision status, self-report was typical.

Among the 12/49 studies reporting on PI-MSM, 9/12 had 400 or fewer PI-MSM participants. For all 12, the association between circumcision status and HIV was a primary outcome of interest, making publication bias more plausible.
